# Gender, acceptability of medically indicated contraception and implications for equitable access to treatment and clinical research in a leishmaniasis-endemic community in Brazil: a mixed-methods study

**DOI:** 10.64898/2026.11.26.26369048

**Authors:** Luiz Henrique Guimarães, Polyana Valente, Ana Flávia Nery, Alex Lago, Clara Cristina de Aguiar, Laura da Silva, Léo Persoz, Fabiana Alves, Diogo Galvão, Denise Pimenta, Gláucia Cota

## Abstract

Requirements for contraception during treatment with potentially teratogenic medicines may create additional barriers to equitable access to care and participation in clinical research. These barriers may be particularly relevant among neglected and underserved populations affected by endemic diseases, where gender inequalities, socioeconomic vulnerability, and limitations in health services often coexist. We examined the individual, social and structural factors shaping the acceptability of medically indicated contraception and their implications for equitable access to leishmaniasis treatment and clinical research.

We conducted an explanatory sequential mixed-methods study, based on community-based participatory research, in rural areas of southern Bahia, Brazil, where cutaneous leishmaniasis is endemic. A cross-sectional survey of 154 women aged 18–49 years at five primary care units assessed willingness to use contraception for future use, if required for oral treatment of cutaneous leishmaniasis and if required to join a clinical trial; the first two answers were compared with an exact McNemar test in the 118 women at risk of pregnancy in the future (not permanently sterile, including pregnant women), women whose answers differed between the two scenarios were described, and the trial scenario was exploratory. The qualitative phase comprised 27 in-depth interviews with survey participants and three focus groups (women community leaders, men and health professionals), analyzed by reflexive thematic analysis. The two datasets were integrated in a joint display and discussed with a community advisory board. Participants lived with marked socioeconomic vulnerability (92.1% in households with up to two minimum wages; 87.0% receiving a government cash transfer). Acceptability of contraception for leishmaniasis treatment was high (129 of 154 women, 83.8%). Among the 118 women at risk of pregnancy in the future (not permanently sterile, including 9 pregnant women), willingness for treatment was not statistically different from future intention to use contraception (87.3% vs 83.9%; difference 3.4 percentage points, 95% confidence interval −4.7 to 11.5; exact McNemar p = 0.54). Answers differed between scenarios for 24 women (20.3%): 14 were willing for treatment only, and 10 intended to use contraception in future but were not willing if it were required for treatment; all 10 had children (median age 32 years). A clinical trial requiring contraception was acceptable to 107 women (69.5%) in an exploratory scenario. Most women reported deciding about contraception alone (61.7%), yet 13.0% had been pressured to become pregnant. Qualitative findings revealed persistent barriers related to gender norms and power struggles in family planning, concerns about contraceptive adverse effects and misinformation, limited acess to health services among younger women, skepticism and trust issues towards research institutions. By integrating the two data types, we observed that acceptability depended less on contraception itself rather than on the context in which it was offered, with convergence on high but conditional acceptability and divergence between reported autonomy and the gender-related constraints described in interviews.

Equitable access to new treatments which require contraception may need more than contraceptive availability. Gender-sensitive counselling, community engagement, reliable information, strengthened health-system pathways and sustained relationships of trust should be incorporated into treatment implementation and clinical research.

## Introduction

Cutaneous leishmaniasis (CL) is a sandfly-borne parasitic disease with an estimated 600,000 to 1 million people become infected annually, mostly in the Americas, Africa, the Middle East and Asia. Brazil is among the countries with the most people affected in the Americas and, in Bahia, particularly in the Corte de Pedra region, transmission has persisted for decades among rural populations with limited access to specialised care. Its impact extends beyond clinical manifestations to psychological distress, stigma, economic loss and reduced quality of life [1–3]. As with other neglected tropical diseases, the burden falls most heavily on communities with high socioeconomic vulnerability and weak health services.

Treatment remains challenging. Pentavalent antimonials require prolonged parenteral courses, cause frequent adverse events and painful injections, and raise fetal safety concerns during pregnancy, while oral options such as miltefosine are potentially teratogenic. Effective contraception is therefore required during treatment and for a period afterwards, adding complexity to treatment access, especially for women of reproductive potential [4–6]. The same requirement affects who can take part in research: concern about pregnancy during studies has historically led to the exclusion or underrepresentation of women, leaving gaps in evidence on efficacy, safety and acceptability, particularly for neglected tropical diseases [7–12]. If new treatments are developed in populations from which women were excluded, or if the conditions attached to treatment are difficult for women to meet, existing inequalities in access to care may deepen. social Brazil, contraceptive use is widespread (over 75% among women in unions), but inequalities by income, education, residence and access persist, and fear of adverse effects, misinformation, religious beliefs, gender norms and limited male support with family planning continue to influence method choice and continuity [13–15]. How these factors operate when contraception is not only a personal choice but also a medical condition for receiving treatment has rarely been examined, and empirical evidence from leishmaniasis-endemic areas on the individual, relational, community and structural social factors involved is scarce [7,8,16].

We conducted a community-based, mixed-methods study to examine the acceptability of medically indicated contraception among women of reproductive age in CL-endemic areas of southern Bahia, and its implications for equitable access to treatment and participation in clinical research. Our central question was whether the contraception requirement itself, or the social and health-system contexts surrounding it, is the main obstacle. A mixed-methods design was chosen because the survey could quantify willingness to use contraception in different scenarios, while the qualitative phase aimed to describe and comprehend the reasons, conditions and gender dynamics behind those responses. The specific objectives were to estimate willingness to use contraception for future family planning, when required for oral treatment of CL, and when required for participation in a clinical trial; to compare willingness between the first two scenarios in women at risk of pregnancy in the future, as a secondary, exploratory objective; to comprehend the demographic and social profile of the women whose answers differed between these scenarios; to explore the perceptions, motivations and concerns underlying these responses; and to identify, with the community, recommendations for more inclusive leishmaniasis care and research. We found that acceptability was high but conditional, and that it depended less on contraception itself than on the contexts in which it was offered.

## Materials and methods

### Study design

We conducted an explanatory sequential mixed-methods study (quantitative followed by qualitative) based on the principles of community-based participatory research (Fig 1) [17,18]. While the components were integrated during interpretation, the quantitative findings informed the design and focus of the qualitative inquiry. The survey characterised contraceptive practices and willingness to use contraception; the qualitative phase explored the experiences and social contexts underlying these findings. Quantitative results informed the interview and focus group guides. A community advisory board (CAB) of community representatives and regional health professionals reviewed the instruments, discussed the results after each study phase (quantitative and qualitative parts) and co-developed recommendations. Reporting followed the STROBE (survey), SRQR (qualitative component) and GRAMMS (mixed methods) guidelines; the checklists are provided in S1 Checklist.

**Fig 1.**
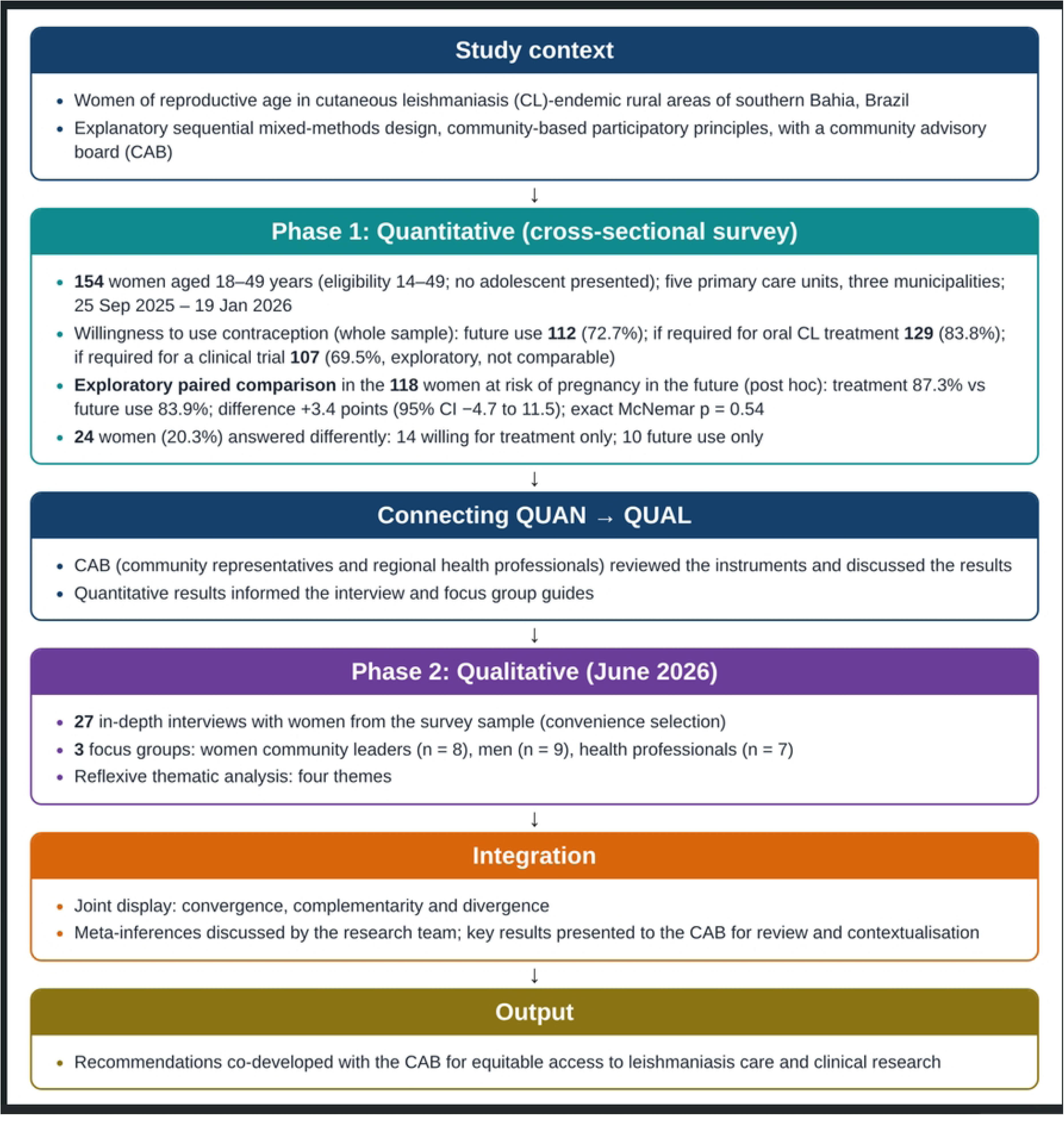
Design of the explanatory sequential mixed-methods study in cutaneous leishmaniasis (CL)-endemic areas of southern Bahia, Brazil. CL, cutaneous leishmaniasis; QUAN, quantitative; QUAL, qualitative. This figure was created by the authors with the assistance of generative artificial intelligence (Claude, Anthropic) and subsequently reviewed and edited by the authors for accuracy and content.

### Setting and participants

The study was conducted in rural communities of three CL-endemic municipalities of southern Bahia (Presidente Tancredo Neves, Teolândia and Wenceslau Guimarães). The survey took place at five primary care units (Corte de Pedra, Alto Alegre, Nova Esperança, São Paulinho and Cocão). Women aged 14–49 years were eligible regardless of contraceptive use, pregnancy or parity; women whom the research team judged unable to answer the questionnaire for clinical reasons were excluded. For adolescents (14–17 years), an assent form and guardian consent were prepared, but no adolescent presented for interview, so they were not used. Women were recruited consecutively during unit visits until the planned sample was reached. The survey was conducted between 25 September 2025 and 19 January 2026. The number of women approached and the reasons for non-participation were not systematically recorded.

### Quantitative phase

The sample size was calculated for the paired comparison of future contraceptive intention and willingness if required for treatment, assuming a baseline proportion of 61.2% (modern contraceptive use among nulliparous women in the region [14]), a 20% relative increase, 95% confidence and 80% power; adding 10% for losses gave 154 women. Trained interviewers administered a structured questionnaire in person, in a private setting and in a standard question order, using KoboToolbox. It covered sociodemographic characteristics; knowledge and experiences of CL; knowledge and use of contraceptive methods; side-effect experiences and concerns; autonomy and decision-making; future intention to use contraception; willingness to use contraception if required for CL treatment; and willingness to participate in a clinical trial. The questionnaire was developed by the research team, administered in Portuguese and piloted with 15 women. The willingness items used a four-point scale (“highly probable”, “probable”, “improbable”, “highly improbable”).

#### Variables and indicators

The outcomes were willingness to use contraception in three scenarios: for future use (future intention), if required for oral CL treatment, and if required to join a clinical trial. Each was measured on a four-point scale (“highly probable”, “probable”, “improbable”, “highly improbable”) and dichotomised into high (“highly probable” or “probable”) and low (“improbable” or “highly improbable”) willingness. Future intention is not applicable to women who are permanently sterile or whose partner has had a vasectomy. The population for comparing future intention with willingness for treatment was therefore the women at risk of pregnancy in the future (n = 118), defined as the 154 participants minus 36 women who were permanently sterile (tubal ligation, 33; hysterectomy, 2) or had a partner with vasectomy (1). The 9 currently pregnant women were kept in this group because they remain at risk of pregnancy in the future and a contraception requirement would apply to them after delivery. Women whose answers differed between the two scenarios (discordant) were classified by direction: high future intention but not willing for treatment (10 women), and low future intention but willing for treatment (14 women). A third group of 14 non-pregnant women who were unwilling to use contraception for treatment (nine of those 10 and five unwilling in both scenarios) was also defined. The clinical trial scenario combines two elements, the use of contraception and participation in a clinical trial, so it cannot be compared with future contraceptive intention, which captures only the former. It is reported descriptively and as exploratory.

#### Sociodemographic, clinical and contraceptive variables were analysed as collected. Missing data are described under Statistical analysis

Analyses was done in SPSS Statistics for Windows, version 23.0 (SPSS Inc., Chicago, Ill., USA). Categorical variables are reported as absolute numbers and percentages (denominator N = 154 unless stated) and continuous variables as means and standard deviations (SD). For the paired comparison in the 118 women at risk of pregnancy in the future, future intention and willingness for treatment were compared with the exact McNemar test (two-sided; p < 0.05 was considered statistically significant). The matched odds ratio (OR) and its exact 95% confidence interval (CI) were calculated from discordant pairs, and the paired difference (in absolute terms) with a Wald 95% CI. The analysis was descriptive and paired. No multiple-comparison correction was needed because a single inferential test was performed. The sample size was calculated for the whole sample, and the restriction to the 118 women was decided after exploring the data (post hoc) and was not prespecified. The comparison is therefore exploratory and secondary to the qualitative findings. It is reported to aid interpretation, not as the test of a main hypothesis. The three groups of women defined above were described with frequencies, means and medians (summary statistics only). Because of the small numbers, no inferential comparison with other women was made.

#### Missing data

Fifteen participants (9.7%) did not respond to each of the questions on monthly household income and main source of income, and one participant did not report education. The “I don’t know” option was selected by 21.4% (n = 33) for transmission and symptoms, 20.1% (n = 31) for women’s susceptibility, 12.3% (n = 19) for prevention and 7.1% (n = 11) for person-to-person transmission. Lower rates were observed for psychological impact (2.6%), treatment knowledge (2.0%) and future disease risk (0.7%). All other variables were complete, and analyses used available data.

### Qualitative phase

During February 2026, we conducted 27 in-depth interviews with women from the survey sample, selected by convenience according to availability and willingness (three of them were among the 14 non-pregnant women who were unwilling to use contraception for treatment). Afterwards, we conducted three focus groups with women community leaders, men from the community and health professionals to obtain complementary perspectives.

Semi-structured guides (S2 Appendix) were developed from the study objectives, the literature, the quantitative results and CAB inputs and were piloted and refined using five pilot initial interviews, which also served to align interviewing techniques across interviewers. Interviews took place at health facilities or in participants’ homes, according to their preference, in the original language of participants, lasted a median of 40 minutes and were conducted by one of three trained female researchers (a historian, an anthropologist and a journalist) with experience in interviewing and community-based research, who had no prior relationship with the participants and were not residents of the study communities. All interviews and focus groups were audio-recorded with consent and transcribed using Transcriptor software. The transcripts were subsequently reviewed and validated by the researchers responsible for conducting the interviews and focus groups, allowing them to check transcription accuracy and incorporate relevant contextual information from field observations. Quotations were translated from Portuguese into English by a bilingual member of the research team and reviewed for semantic accuracy. Audio files, transcripts and analytical datasets were stored on password-protected servers with access restricted to authorised study personnel. Data were analysed using a thematic approach informed by the reflexive thematic analysis proposed by Braun and Clarke [19].

The validated transcripts were analysed by two researchers involved in the fieldwork. Following initial familiarisation with the data through repeated readings, the material was organised and coded using Microsoft Excel. Initial codes were informed by the study objectives and semi-structured interview guides and were progressively refined to incorporate additional analytical categories developed through engagement with the empirical material. Interviews, focus groups and field notes were examined comparatively to identify recurring patterns, divergent perspectives and context-specific experiences. Preliminary findings and thematic interpretations were discussed with the broader research team and subsequently presented to the community advisory board (CAB). Four themes were ultimately defined.

### Integration and interpretive framework

Integration occurred at the design, methods and interpretation levels [18]. At the design level, the quantitative results informed the interview and focus group guides. At the interpretation level, we built a joint display that set the quantitative results alongside the qualitative themes for analytical dimension. Each dimension was classified as convergence (confirmation), complementarity (expansion) or divergence (discordance) [18]. From each row, the research team drew a meta-inference, defined as a conclusion generated by integrating the quantitative and qualitative findings [20]. The meta-inferences are the “integrated interpretation” column of the joint display and are developed in the Results (Integration of quantitative and qualitative findings). Those that went beyond what the data show, such as the explanation of the differing answers in the two scenarios, are labelled as interpretations in the text [20]. Key results were presented to the CAB for review and contextualisation.

In interpreting the meta-inferences, we adopted a health equity lens [21], which views differences in health and access to care as shaped by social, economic and structural conditions and by power inequalities, not only by individual choice. Within this lens we distinguished individual (for example, concerns about adverse effects), relational (for example, partner and family influence) and structural (for example, socioeconomic conditions, health services and trust in institutions) factors. The structural level draws on the concept of structural vulnerability [22].

### Ethics statement

This study was conducted in accordance with Brazilian National Health Council Resolution No. 466/2012, which establishes the ethical and regulatory requirements for research involving human participants in Brazil and recognises the Declaration of Helsinki among its international ethical foundations. The protocol was approved by the Research Ethics Committee of the Federal University of Recôncavo da Bahia (CAAE 86549124.0.0000.0056; Opinion 7.720.342, 22 July 2025) and by the municipal health secretariats. This observational study is not a clinical trial; registration is not mandatory, and the study was registered voluntarily and retrospectively in the Brazilian Registry of Clinical Trials (ReBEC) (RBR-4mccfgk; Universal Trial Number U1111-1329-1868) on 11 March 2026, after ethical approval and after the survey had started. An assent form for adolescents was developed but not used. All participants provided written informed consent, including consent to audio recording and the use of anonymised testimonies.

## Results

### Participants and socioeconomic context

We enrolled 154 women aged 18–49 years (mean 32.7, SD 8.2; eligibility started at 14 years of age, but no adolescent presented for interview) at five primary healthcare facilities in three municipalities of southern Bahia; all completed the questionnaire (Table 1). Most identified as Pardo or Black (95.5%), lived with a partner (85.1%) and in rural areas (76.0%; 86.4% had a direct connection to rural settings). Catholic (53.9%) and Evangelical (31.8%) affiliations predominated, and secondary education was the most frequent level (61.0%). Socioeconomic vulnerability was marked: 92.1% of households lived on up to two minimum wages, 87.0% received Bolsa Família, a government-funded social protection programme for low-income households, and only 9.7% of the women held formal employment.

**Table 1.** Sociodemographic and socioeconomic characteristics of the participants (N = 154)

| **Characteristic** | **n (%)** |
|---|---|
| Age, years, mean (SD) | 32.7 (8.2) |
| Ethnicity/skin colour: Pardo / Black / other | 74 (48.1) / 73 (47.4) / 7 (4.5) |
| Living with a partner | 131 (85.1) |
| Currently pregnant | 9 (5.8) |
| Number of children, mean (SD) | 1.93 (1.37) |
| Residence: rural / urban / both | 117 (76.0) / 21 (13.6) / 16 (10.4) |
| Education: primary / secondary / higher | 41 (26.6) / 94 (61.0) / 18 (11.7) |
| Religion: Catholic / Evangelical / none / other | 83 (53.9) / 49 (31.8) / 18 (11.7) / 4 (2.6) |
| Occupation: homemaker / self-employed / informal service / formal employment / student | 59 (38.3) / 47 (30.5) / 27 (17.5) / 15 (9.7) / 6 (3.9) |
| Main income from government programmes | 75 (54.0) |
| Household receives Bolsa Família | 134 (87.0) |
| Household income up to two minimum wages | 128/139 (92.1) |

Education: one participant (0.6%) did not report her level; “higher” includes partial or complete attendance. Ethnicity “other”: White 3, Asian 3, Indigenous 1. SD, standard deviation. Income rows are calculated on the 139 women who answered (15 did not respond).

### Knowledge, experience and perceptions of leishmaniasis

Awareness of CL was high (Table 2): 93.5% had heard of it, 89.6% knew someone affected, and 50 of the 144 women aware of it (34.7%) had had CL. Health professionals were the main source of information (33.1%). Knowledge was incomplete: about 70% named the vector, 11% gave answers reflecting misconceptions (free text: ticks, fleas, worms, contaminated blood, contact with wounds), while 14.3% believed in person-to-person transmission. A quarter (26.6%) reported no preventive measure. CL caused fear or psychological distress in 72.7%; the main concerns about treatment were injection pain (49.4%), not being cured (29.2%), lost income or extra expenses (24.7%) and daily travel for injections (24.7%), whereas stigma was rarely cited (3.3%).

**Table 2.** Knowledge, experience and perceptions of cutaneous leishmaniasis (CL) (N = 154)

| **Item** | **n (%)** | **Free-text responses** |
|---|---|---|
| Has heard of CL | 144 (93.5) |  |
| Knows someone affected | 138 (89.6) |  |
| Has had CL (among 144 aware) | 50 (34.7) |  |
| Information from health professionals / family or neighbours / school | 51 (33.1) / 23 (14.9) / 10 (6.5) |  |
| Believes CL is transmitted person to person: yes / no / unsure | 14.3% / 72.1% / 7.1% |  |
| Recognises skin lesion as a symptom | 89 (57.8) | Enlarged lymph nodes (“íngua”) frequently cited |
| Prevention: insect repellent / no measure | 45 (29.2) / 41 (26.6) | Cleaning, avoiding standing water; incorrect measures (vaccine, sunscreen, incense) |
| CL causes fear or psychological distress | 112 (72.7) |  |
| Concerns about CL treatment |  |  |
| Injection pain | 76 (49.4) | Side effects, body pain, long treatment and number of injections, effect on daily activities and school, fear of death or sequelae (n = 46) |
| Not being cured | 45 (29.2) |  |
| Loss of income or extra expenses | 38 (24.7) |  |
| Daily travel to the health facility | 38 (24.7) |  |
| Cost of consultations and treatment | 37 (24.0) |  |
| Anxiety or depression / scarring | 35 (22.7) / 32 (20.8) |  |
| Stigma / no concerns | 5 (3.3) / 13 (8.4) |  |

Percentages are calculated on N = 154 unless stated; multiple responses were allowed.

### Contraceptive use, knowledge and experiences

Contraceptive experience was extensive (Table 3): 94.1% had ever used a method and 56.5% were current users, mainly of pills (57.1%) and injectables (three-monthly 48.1%, monthly 32.5%). Pills (97.4%) and injectables (95.5%) were the best-known methods, compared with implants (39.6%) and emergency contraception (33.1%). Of the 67 non-users, 45 were not at risk of pregnancy (9: pregnant; 36: permanent contraceptive method, including 1 case in which the partner had undergone vasectomy); among the 22 at risk, the following reasons were informed (more than one reason could be informed): no specific reason (7), sexual inactivity (6) and wanting more children (3). Only four of the 70 women who gave reasons for non-use opposed contraception, and two cited lack of access. Weight gain (61.0%) and nausea or vomiting (53.3%) were the side effects most often experienced or heard about and the main concerns for future use (41.6% and 29.9%); free-text answers added thrombosis, cardiovascular risk and pregnancy despite contraception. Most women decided on their own (61.7%) or jointly with their partner (37.0%), yet 13.0% reported having been pressured to become pregnant against their wishes.

**Table 3.** Contraceptive use, knowledge, side effects and decision-making.

| Item | n (%) | Free-text responses |
|---|---|---|
| Ever used contraception / currently using | 145 (94.1) / 87 (56.5) |  |
| Non-users not at risk of pregnancy (of 67) | 45 (67.2)# |  |
| Reasons for non-use: permanent method/ partner infertility / pregnant / fear of side effects / not in favour/intention to get pregnant/ don’t know/others | 33 /1/ 9 / 10 / 4/3/6/4 | Others: one medical restriction (epilepsy), one cost, two lack of local access |
| **Methods ever used** |  |  |
| Daily pill / three-monthly injectable / monthly injectable | 88 (57.1) / 74 (48.1) / 50 (32.5) |  |
| Male condom / sterilisation or partner vasectomy/ emergency contraception / IUD | 56 (36.4) / 36 (23.4) / 15 (9.7) / 6 (3.9) |  |
| **Methods known** |  |  |
| Pill / injectable / IUD / male condom | 150 (97.4) / 147 (95.5) / 105 (68.2) / 102 (66.2) |  |
| Implant / emergency contraception | 61 (39.6) / 51 (33.1) |  |
| **Side effects experienced or heard about** |  |  |
| Weight gain / nausea or vomiting | 94 (61.0) / 82 (53.3) | Dizziness, oedema, thrombosis, anxiety, injection-site pain |
| Cramps / headache / acne | 44 (28.6) / 43 (27.9) / 36 (23.4) |  |
| Irregular / heavier / reduced bleeding | 35 (22.7) / 34 (22.1) / 30 (19.5) |  |
| **Side effects of concern for future use** |  |  |
| Weight gain / nausea or vomiting / headache / heavier bleeding | 64 (41.6) / 46 (29.9) / 30 (19.5) / 29 (18.8) | Thrombosis, stroke with hypertension, pregnancy despite contraception |
| **Decision-making** |  |  |
| Decides herself / jointly with partner / other person | 95 (61.7) / 57 (37.0) / 2 (1.3) |  |
| Joint decision (n = 57): her opinion equal / more / less important | 34 (59.6) / 20 (35.1) / 3 (5.3) |  |
| Ever pressured to become pregnant against her wishes | 20 (13.0) |  |

Percentages are calculated on N = 154 unless stated; multiple responses were allowed. Permanent methods: tubal ligation (n = 33), hysterectomy (n = 2) and partner vasectomy (n = 1). IUD, intrauterine device. # 9 pregnant, 36 sterilised

### Willingness to use contraception

Willingness was high in all three scenarios (Table 4): 72.7% reported a high likelihood of future contraceptive use, 83.8% were willing to use contraception if required for oral CL treatment, and 69.5% were willing to join a clinical trial requiring contraception (to prevent possible fetal risk). Most would accept contraception for more than six months (68.2% in the treatment scenario and 62.6% in the trial scenario), and the daily pill was the preferred method in both scenarios (39.0% and 29.9%), followed by the three-monthly injectable (24.0%). The clinical trial scenario combined two elements, contraception and participation in a trial, and is presented descriptively; it is not directly comparable with future contraceptive intention.

**Table 4.** Willingness to use contraception under three scenarios (N = 154)

|  | **Future use for family planning** | **Future use as a requirement for CL treatment** | **Clinical trial participation** |
|---|---|---|---|
| High willingness (highly probable + probable) | 112 (72.7) | 129 (83.8) | 107 (69.5) |
| Highly probable | 70 (45.5) | 94 (61.0) | 64 (41.6) |
| Probable | 42 (27.3) | 35 (22.7) | 43 (27.9) |
| Low willingness (improbable + highly improbable) | 42 (27.3) | 25 (16.2) | 47 (30.5) |
| Would accept contraception for more than 6 months | – | 88/129*(68.2) | 67/107* (62.6) |
| Preferred method: pill / three-monthly injectable / IUD | – | 60 (48.4) / 37 (29.8) / 27 (21.7) among 124 women | 46 (43.8) / 37 (35.2) / 22 (20.9) among 124 women |

Values are n (%) of N = 154, except when denominator is informed\*. IUD, intrauterine device. The clinical trial scenario combines contraception and trial participation and is not comparable with future use.

In the 118 women at risk of pregnancy in the future (Table 5), 103 (87.3%) were willing to use contraception if required for oral CL treatment and 99 (83.9%) reported high future intention for family planning (absolute difference 3.4 percentage points, 95% CI −4.7 to 11.5). Eighty-nine women were willing in both scenarios, 5 in neither, 14 only for treatment and 10 only for family planning future use (exact McNemar p = 0.54; matched OR 1.40, 95% CI 0.58–3.52). The interval is compatible with no difference and with a difference of up to about 11 percentage points in favour of willingness to use contraception if required for CL treatment, so the comparison is imprecise. Overall, 24 women (20.3%) answered differently in the two scenarios, in both directions. The 36 permanently sterile women were excluded from this comparison because future intention does not apply to them.

**Table 5.** Paired comparison of willingness to use contraception for oral CL treatment and for future family planning in the 118 women at risk of pregnancy in the future (including 9 pregnant women)

| Women at risk of pregnancy in the future (n = 118) | **Future family planning intention: high** | **Future family planning intention: low** | **Total** |
|---|---|---|---|
| Willing to use for oral CL treatment | 89 | 14 | 103 (87.3%) |
| Not willing to use for CL treatment | 10 | 5 | 15 (12.7%) |
| Total | 99 (83.9%) | 19 (16.1%) | 118 |

Matched OR = 14/10 = 1.40 (exact 95% CI 0.58–3.52); exact McNemar p = 0.54; Difference (in absolute terms) +3.4 percentage points (Wald 95% CI −4.7 to 11.5). High willingness: “highly probable” or “probable”. CL, cutaneous leishmaniasis; OR, odds ratio; CI, confidence interval. The 118 women are those not permanently sterile and include 9 pregnant women, who remain at risk of pregnancy in the future

### Women whose answers differed between the two scenarios

Of the 24 women with discordant answers (20.3% of the 118), 14 (58.3% of the 24; 11.9% of the 118) were willing to use contraception if required for treatment but had low future intention, and 10 (41.7% of the 24; 8.5% of the 118) had high future intention but were not willing to use contraception if it were required for oral CL treatment. These 10 women were aged 20 to 48 years (mean 34.7, median 32; whole sample, mean 32.7) and all had children (mean 2.5; whole sample, 1.93). Two were past users of contraception, and three of nine had CL, similar to the whole sample, so they were not women unfamiliar with contraception or with CL. Because they intended to use contraception in the future, their refusal appears to relate to the requirement more than to contraception itself. These 10 women overlap with the 14 non-pregnant women who were unwilling to use contraception for treatment (nine of the 10, plus five unwilling in both scenarios); only three of these 14 women took part in the individual interviews. The five who were unwilling in both scenarios (aged 29 to 36 years, including the only childless woman) did not differ markedly in age or parity from the nine with high future intention (aged 21 to 48). The 14 women with the opposite pattern (willing for CL treatment, low future intention) were of similar age (18 to 47 years; median 34) but had fewer children (mean 1.7 vs 2.5) and at least four were childless, compared with none of the 10. These figures rest on the number of very few women, were not compared statistically with those of other women, and do not show the reasons for their answers.

### Qualitative findings

The qualitative phase included 27 in-depth interviews with women of childbearing potential and three focus groups with women community leaders (8 participants), men (9) and health professionals (7). Thematic analysis identified four themes: (1) leishmaniasis as an experience of physical, emotional and social suffering; (2) navigating contraception: autonomy, gender-related constraints and perceived risks; (3) medically indicated contraception perceived differently from contraception for family planning; and (4) trust and information prerequisites for participation in clinical research. The integration of these themes with the survey results is presented in the next section.

#### Theme 1. Leishmaniasis as an experience of physical, emotional and social suffering

This theme combined four groups of codes: pain and treatment burden; scars and stigma; gender and caregiving responsibilities; and collective memory of treatment. Participants described the pain of injectable treatment, permanent scars, social stigma and effects on self-esteem. Women reported difficulty performing domestic tasks, caring for children and keeping up laubor activities during treatment. The number of injections and the associated pain were emphasised in the focus group with women community leaders:

> “Sixty injections were not enough for the lesion to heal. So I had to repeat the treatment, reaching 120 injections. One hurt in my leg, another in my abdomen. My goodness, no one has experienced that pain… I almost died. What kind of pain is that?” (Woman community leader, focus group)

Scars were linked to shame, social stigma and restrictions on social interaction which persisted long after physical recovery:

> “We often think about those people who are left with a visible scar, who sometimes stop doing certain things because of that scar, you know, which remains forever. And sometimes people carry it in their souls. And those who carry it in their souls suffer a lot.” (Woman community leader, focus group)

Lived experiences and discourses varied. Some participants described less disruptive treatment experiences or limited long-term consequences, and others accepted treatment as a necessary part of recovery despite the discomfort. The collective memory of conventional treatment was largely negative and often described as traumatic, built from individual accounts, family experiences and narratives shared in the focus groups. A minority expressed confidence in existing treatments after seeing relatives or community members recover. Participants mainly welcomed the prospect of an oral treatment as a way to reduce treatment burden and avoid repeating familiar painful experiences.

#### Theme 2. Navigating contraception: autonomy, gender-related constraints and perceived risks

Most women reported autonomy in contraceptive decisions, but also described machismo, unequal power dynamic, the need to negotiate with partners and limited male participation in contraception, particularly regarding condom use. Decisions were often shaped by partners’ expectations and attitudes. Other women described supportive relationships with joint decisions, and some men regarded pregnancy prevention as a shared responsibility. In the focus group with healthcare professionals, participants described vulnerabilities for women with limited acess to health services, particularly younger women, who faced restrictions from partners or family on contraceptive use and access to care; adolescent pregnancy and vulnerability to sexually transmitted infections were also mentioned.

Concerns about adverse effects, contraceptive failure and long-term health consequences recurred. The intrauterine device (IUD) and hormonal methods were both desired and feared. Women who had undergone tubal ligation, or who considered it, often described it as a definitive solution which removed uncertainty about effectiveness. Among rural women, concerns about the IUD were sometimes linked to the physical demands of agricultural work, whereas concerns about hormonal methods centred on menstrual changes, weight gain and general health.

> “Some women are afraid of the contraceptive because they say it makes them feel sick, gain weight, or causes other problems. Sometimes they prefer not to use it because they are worried about the side effects.” (Health professional, focus group)

These perceptions drew on personal experience and on knowledge circulating in families and communities. Some women reported positive experiences, expressed confidence in the safety and effectiveness of their method of choice and voiced fewer concerns about side effects. At the same time, participants highlighted how relationship dynamics and partner approval could represent substantial barriers to contraceptive uptake, and even to seeking care:

> “Here, men still have a lot of control. We have to ask permission for almost everything, even to go to the doctor or take medication.” (Female community leader, focus group)

#### Theme 3. Medically indicated contraception is perceived differently from contraception for family planning

Many women gave different meanings to contraception for family planning and temporary contraception for medical reasons. Preventing treatment-related risks in pregnancy was generally accepted as a legitimate reason for temporary use. Some, however, saw the requirement as adding additional concerns to an already difficult treatment because of concerns about side effects, and accepting contraception for medical reasons did not remove fears of menstrual changes, weight gain or other adverse effects. Other participants did not distinguish between the two situations: women already willing to use contraception said a temporary medical requirement would not change their decision, and others saw it as a routine health recommendation.

Perspectives differed between groups. Healthcare professionals described contraception mainly as a risk-mitigation measure, whereas women and women community leaders spoke from personal experience, highlighted concerns about adverse effects and relationship dynamics.

Men’s attitudes towards treatment and research participation were mostly favourable and mainly geared towards women, but concerns about contraceptive failure and pregnancy during use of an investigational product remained:

> “If it means she won’t have to suffer through those injections anymore, I support anything. But what if the medication fails and she gets pregnant? What happens to the baby’s health?” (Male participant, focus group)

A smaller number of men and women said that adequate information and medical follow-up would reduce these risks.

#### Theme 4. Trust and information as prerequisites for participation in clinical research

Willingness to take part in research was closely tied to trust. Participants valued clear information on risks and benefits, an explanation of how studies work, and close relationships with health professionals and researchers.

> “If the researchers explain everything clearly and stay close to the community, people feel more confident. Trust is built through the relationship they establish with us.” (Woman of childbearing age, in-depth interview)

Community health agents were described as key intermediaries because communities already knew and trusted them, and the participants asked that study results be returned to the community as a sign of respect. Trust was not uniform: some participants worried about uncertainty over investigational treatments, potential risks and unforeseen consequences, and said that researchers had to earn trust through transparency, continued communication and visible commitment rather than rely on institutional affiliation. Most participants understood and accepted contraception as a way to prevent fetal exposure to treatment-related risks. Their willingness to join a trial, however, also depended on trust in the research team, uncertainty about investigational products, family influence and their own appraisal of risks and benefits.

Some were enthusiastic about contributing to the research and to the improvement of treatments, while others remained cautious despite understanding the scientific rationale.

### Integration of quantitative and qualitative findings

We used a joint display (Table 6) to compare the quantitative results with the qualitative themes and to identify convergence (confirmation), complementarity (expansion) and divergence (discordance) between them, and to draw a meta-inference for each analytical dimension (last column of the table).

**Table 6.** Joint display of quantitative and qualitative findings on contraceptive acceptability, leishmaniasis treatment and clinical research.

| **Analytical dimension** | **Quantitative findings (N = 154)** | **Qualitative findings** | **Integrated interpretation (meta-inference)** |
|---|---|---|---|
| Acceptability for oral treatment | 129 (83.8%) high willingness if contraception is required for oral CL treatment (94 [61.0%] highly probable); among 118 women at risk of pregnancy in the future: 103 (87.3%) willing for treatment vs 99 (83.9%) with high future intention, difference +3.4 points (95% CI −4.7 to 11.5), p = 0.54; 24 (20.3%) answered differently | Painful, prolonged injections and negative collective memory; oral treatment welcomed (Theme 1). Medical contraception seen as legitimate, but additional concern for some (Theme 3) | Convergence (confirmation) with elaboration: acceptability is high but conditional on concerns about side effects |
| Contraceptive experience | 145 (94.1%) ever used; 87 (56.5%) current users; 45 of 67 non-users not at risk of pregnancy | Views shaped by personal and community experience; positive and negative accounts; tubal ligation as definitive solution (Theme 2) | Complementarity (expansion): qualitative accounts contextualise use and non-use |
| Women whose answers differed between scenarios | 10 (8.5% of 118) with high future intention but unwilling for treatment (all with children; median age 32); 14 (11.9% of 118) the reverse (similar age, fewer children, at least 4 childless); of 14 non-pregnant women unwilling for treatment, 11 also refused a trial requiring contraception | Only three of the 14 non-pregnant women unwilling for treatment took part in the individual interviews. Other participants distinguished contraception for family planning from temporary medical contraception while some accounts suggested that treatment-related contraceptive requirements could introduce additional concerns (Theme 3)<br>” (Theme 3) | Complementarity (expansion, inferential): answers differed in both directions, consistent with Theme 3; the reasons of the discordant women are largely unknown |
| Side effects and perceived risk | Experienced or heard of: weight gain 94 (61.0%), nausea 82 (53.3%). Feared: weight gain 64 (41.6%), nausea 46 (29.9%) | Fear of adverse effects, failure and long-term harm; IUD and hormonal methods both desired and feared; IUD concerns linked to agricultural work | Convergence (confirmation): concern about side effects present in both components |
| Reproductive autonomy | 95 (61.7%) decide alone; 57 (37.0%) jointly with partner | Autonomy exercised within negotiation with partners, machismo, limited male participation; supportive partners also described; professionals report restrictions on younger women | Divergence (discordance): reported autonomy coexists with gender-related constraints |
| Reproductive pressure | 20 (13.0%) pressured to become pregnant against their wishes | Partner and family influence on reproductive choices | Complementarity (expansion): qualitative data illustrate how power relations operate |
| Treatment burden and stigma | Injection pain 76 (49.4%); fear or distress 112 (72.7%); anxiety or depression 35 (22.7%); lost income 38 (24.7%); daily travel 38 (24.7%); scarring 32 (20.8%); stigma 5 (3.3%) | Pain and number of injections, scars, shame and social judgement, difficulty with domestic work and childcare (Theme 1) | Convergence (confirmation) with elaboration; partial divergence (discordance) for stigma, which the closed-ended item may under-estimate |
| Information and knowledge | 144 (93.5%) had heard of CL; about 21.4% did not know how it is transmitted; health professionals' main source of information, 51 (33.1%) | Clear information on risks and benefits valued; community health agents trusted (Theme 4) | Complementarity (expansion): information needs and trusted channels |
| Clinical trial participation | 107 (69.5%) high willingness; 47 (30.5%) low willingness (exploratory; combines contraception and trial participation, not comparable with future intention) | Trust in research team, uncertainty about investigational products, family influence; men concerned about failure and fetal health (Themes 3 and 4) | Convergence (confirmation) with conditions: willingness does not imply unconditional consent |
| Trust in research | No direct indicator | Trust in community health agents and familiar professionals; return of results to the community (Theme 4) | Qualitative-only insight (expansion) |

CL, cutaneous leishmaniasis; CI, confidence interval; OR, odds ratio. Quantitative values are n (%) of N = 154 unless stated; the difference is in percentage points.

The two components converged on high acceptability of contraception when it is framed as protection during treatment. Overall, 129 women (83.8%) were willing to use contraception to receive an oral treatment, and the qualitative data describe why: the pain and burden of injectable treatment, the scars and the negative collective memory of conventional therapy made an oral alternative attractive (Theme 1). The qualitative findings also show that this acceptability is conditional. Among the 118 women at risk of pregnancy in the future, willingness for treatment was similar to future intention to use contraception (87.3% vs 83.9%), so the treatment framing did not by itself raise acceptability. Answers nevertheless differed between scenarios for one in five women, in both directions (14 willing only for CL treatment, 10 only for future family planning use). The two discordant groups differed in parity, the women willing only for CL treatment including more childless women. This pattern is consistent with Theme 3, in which participants distinguished contraception for family planning from temporary contraception for medical reasons: some accepted the latter as legitimate protection without wanting the former, whereas others, who intended to use contraception, may have perceived treatment-related contraceptive requirements as an additional concern or burden.. The survey cannot tell which explanation applies to the discordant women, and only three of the 14 non-pregnant women unwilling for treatment (nine of whom had high future intention) took part in the individual interviews, so this link is an interpretation and not a finding. Fears of side effects persisted even among women who accepted contraception for medical reasons (Theme 3).

The main divergence concerned reproductive autonomy, a relational factor. Most women reported deciding on contraception alone (61.7%) or jointly with their partner (37.0%), yet 13.0% had been pressured to become pregnant against their wishes, and qualitative findings accounts described machismo, power struggles with partners and limited male participation, especially in condom use. Healthcare professionals also described younger women who faced restrictions from partners or family. Reported autonomy therefore coexisted with gender-related constraints that closed-ended items did not capture. A second divergence concerned stigma: 5 women (3.3%) selected community knowledge of the diagnosis as a concern about treatment, although 32 (20.8%) feared scarring, and qualitative accounts linked visible scars to shame and social stigma.

Concerns about contraceptive adverse effects were consistent across components. Women had wide experience of contraception (145 [94.1%] ever users; 87 [56.5%] current users), but weight gain was experienced or heard of by 94 (61.0%) and feared by 64 (41.6%). The interviews described fear of IUD and hormonal methods alongside the wish for them, and tubal ligation as a definitive solution (33 women, 21.4%, had undergone tubal ligation). Willingness to join a trial requiring contraception was 69.5% (107 women). This scenario combines contraception and participation in a trial, so it cannot be compared with future contraceptive intention and is described narratively, as an exploratory observation. The qualitative data point to reasons beyond contraception: trust in the research team, lack of awareness and confidence in the efficacy and safety of investigational products, family influence and, among men, concern about contraceptive failure and fetal health (Themes 3 and 4). Health professionals were the main source of information on CL (51 women, 33.1%), in line with the central role of professionals and community health agents in building trust, but no quantitative item measured trust directly.

Taken together, the findings suggest that contraceptive acceptability in this setting depends on context and is shaped by treatment expectations, past contraceptive experiences, gender relations and trust, and not only by individual preference. Stated willingness is also hypothetical and may overstate what women will do in practice. The survey did not measure access or equity directly; the equity implications are drawn from the qualitative findings and from the recommendations co-developed with the community advisory board.

### Recommendations co-created with the community advisory board

The participatory analysis of the results allowed recommendations to be aimed at health services, leishmaniasis control programmes and future clinical research (Table 7).

**Table 7.** Recommendations co-developed with the community advisory board for equitable access to leishmaniasis care and clinical research.

| **Domain** | **Recommendations** |
|---|---|
| Strengthening leishmaniasis care | Periodic training of health professionals on the diagnosis, treatment and monitoring of leishmaniasis; strengthening the integration between primary care and the referral centre; definition of regional care flows for case monitoring; expansion of educational actions on leishmaniasis in communities and schools. |
| Expanding access to family planning | Diversification of the supply of contraceptive methods; improvement of contraceptive counselling; strategies for the inclusion of men in reproductive decisions; specific actions for adolescents and women in vulnerable situations. |
| Inclusion of women in clinical research | Accessible educational materials on clinical research; strengthening of ongoing community communication; active participation of the community in all phases of studies; transparency about risks, benefits and research results; use of participatory structures, such as CABs, to support the development and implementation of future clinical research. |

Together, these recommendations point to an integrated approach which combines strengthening health services, promoting reproductive autonomy and equity, community participation and gender-sensitive strategies as elements that may help expand equitable access to leishmaniasis treatment and clinical research involving all communities affected by the disease, not only women of childbearing age.

## Discussion

### Principal findings

In this explanatory sequential mixed-methods study of 154 women in a CL-endemic area of southern Bahia, most women were willing to use contraception if it were required for oral CL treatment (83.8%). Among the 118 women at risk of pregnancy in the future, this was similar to willingness to use contraception for future family planning (87.3% vs 83.9%), although one in five women answered differently in the two scenarios. Ten women (8.5% of the 118) intended to use contraception in future family planning but were not willing to use it if required for CL treatment, and all of them had children. In an exploratory scenario, 69.5% would join a clinical trial requiring contraception; this cannot be compared with future contraceptive intention because it combines contraception and trial participation. For the women who accepted, acceptability depended less on contraception itself than on the gender, socioeconomic and health-system context in which it would be offered; the barriers facing the women who were unwilling are only partly identified, as only three of the 14 non-pregnant women unwilling for treatment took part in the individual interviews. The qualitative phase and the integrated analysis indicate that these willingness figures were conditional.

### Contraception as a requirement: acceptable, but conditional

The comparison between the treatment and future-use scenarios should be read cautiously. Among the 118 women at risk of pregnancy in the future, willingness in the two scenarios was similar (87.3% vs 83.9%; difference 3.4 points, 95% CI −4.7 to 11.5; p = 0.54), so these data do not show that framing contraception as a treatment requirement raised acceptability, although the interval does not exclude a difference of up to about 11 points. More informative is that one in five women answered differently, in both directions. Ten women intended to use contraception in future family planning but were not willing to use it as a condition of CL treatment. They were not unfamiliar with contraception or with CL, all had children, and their median age (32 years) was close to that of the sample; this pattern raises the possibility that requiring contraception as a condition of treatment may introduce concerns beyond those associated with contraceptive use itself. However, the available data do not allow us to establish the reasons for these women’s responses. Among the 14 non-pregnant women unwilling to use contraception for CL treatment, the five who also had low future intention for family planning were of similar age and parity to the nine who intended to use contraception (ages 29–36 vs 21–48; the only childless woman was among the five), so there was no clear distinct profile. Fourteen other women showed the opposite pattern, willing to use contraception for CL treatment but with low future intention for family planning. They were of similar age but less parous (mean 1.7 children; at least four childless), so the two discordant groups differed mainly in parity: refusal of the requirement was concentrated among women with children and established use, whereas acceptance without future plans included more childless women. This fits the distinction between contraception for family planning and temporary contraception for medical reasons (Theme 3) and may reflect no current need or plan, but their reasons were not asked. With so few women the profiles are descriptive and were not compared with those of other women. Temporary contraception linked to treatment was seen by many women as legitimate protection, in line with the view that acceptability depends on perceived effectiveness and on the value attributed to the expected benefit [23]. However, recognising the medical necessity of contraception did not mean unconditional acceptance: the requirement could introduce additional concerns related to contraceptive use, particularly regarding weight gain, menstrual changes and other potential adverse effects.. The painful, prolonged and socially stigmatising experience of conventional treatment, consistent with the psychosocial burden of CL reported elsewhere [24–26], probably increased receptivity to an oral treatment, which women saw as a way to avoid repeating painful experiences.

### Relational and structural factors shaping access

Most women reported deciding on contraception alone (61.7%) or jointly with a partner (37.0%), yet 13.0% had been pressured to become pregnant, and the qualitative accounts described machismo, negotiation with partners, limited male participation and the need to ask permission even to seek care, as found in other rural and socially vulnerable settings [13,27–29]. These relational constraints are a gender equity issue that a contraception requirement may amplify: a woman who must negotiate with a partner to use a method, or to attend a health service, may be unable to meet the conditions attached to treatment even when she is willing. The autonomy reported by participants may also not reflect women in more vulnerable circumstances, such as younger women facing partner or family restrictions, who were probably under-represented, as health professionals noted.

Contraceptive acceptability also cannot be understood from access and information alone. Most participants knew and had used contraceptive methods (94.1% ever users) and few non-users cited lack of access, but concerns about side effects, shaped by personal experience and shared narratives, remained, as reported in other low- and middle-income settings [28,30,31]. The broader socioeconomic context is relevant to how such a requirement would be experienced: most households lived on up to two minimum wages and depended on social protection, few women had formal employment, and treatment already implied lost income, extra expenses and daily travel for about a quarter of the women. In this context, an additional requirement that depends on repeated contact with health services, on the availability of a range of methods and on reliable counselling may be easier to meet for some women than for others. Research teams and services should therefore address both concerns and positive experiences with contraception in counselling, rather than only providing technical information.

### Trust and the inclusion of women in clinical research

These findings add to the evidence that concern about pregnancy during research has led to the exclusion of women of reproductive age from clinical trials [10–12]. Willingness to join a trial requiring contraception was 69.5% (an exploratory observation that cannot be compared with future contraceptive intention, because it combines contraception and trial participation), and the qualitative data show that it depended on trust in the research team, uncertainty about investigational products and family influence, not only on contraception. Trust emerged as a central condition for participation: participants valued clear information on risks and benefits, the role of community health agents and the return of results, in line with the literature on community engagement in research [32–35]. Participatory structures such as community advisory boards can support these processes and ensure that women’s concerns shape how contraception requirements are designed and explained.

### Implications for equitable access to treatment and research

Taken together, our results suggest that making contraception available is necessary but not sufficient. The recommendations co-developed with the CAB (Table 7) translate this into practice: diversify the supply of methods and improve counselling; involve men in reproductive decisions and attend to adolescents (who were not reached by our survey) and women in vulnerable situations; strengthen the pathways between primary care and the referral centre; and ensure transparent, continuous communication with the community, including the return of results. When new oral treatments for leishmaniasis are implemented, or evaluated in trials, these elements should be built into the programme from the start so that the requirement for contraception does not itself become a source of inequity.

### Limitations

Recruitment through health facilities may have favored women with greater access to services and stronger links with providers. Health professionals noted that younger women with early union, partner-imposed restrictions and greater vulnerability may have been under-represented. Although eligibility included girls aged 14–17 years, none presented for interview, so the findings apply to women aged 18–49 and cannot be extended to adolescents.

The interviewees were drawn from the survey sample by convenience, so selection bias may have affected both components. In particular, only three of the 14 non-pregnant women unwilling to use contraception for treatment took part in the individual interviews, so their perspective is under-represented in the qualitative data.

Willingness was hypothetical and may overstate actual behavior, and the four-point items were dichotomised. Responses may also have been affected by social desirability and interviewer effects. No quantitative item measured trust or health-service access directly, so these dimensions rest mainly on the qualitative data. Clustering by primary care unit was not accounted for in the analysis. The comparison between future intention and willingness for treatment was post hoc and restricted to women at risk of pregnancy in the future, and it rests on few discordant pairs (24), so it is imprecise. The same small numbers limit the descriptions of the women whose answers differed between scenarios, which are not supported by comparison groups or by the women’s own reasons. The clinical trial scenario combines contraception and trial participation, so it cannot be compared with future contraceptive intention and is reported only as an exploratory observation. The study covered a single region, which limits generalisability.

## Conclusions

Most women in this CL-endemic area said they would use contraception if it were required for an oral treatment (83.8%), and 69.5% would join a clinical trial requiring it (an exploratory observation, since this scenario combines contraception and trial participation). Among the 118 women at risk of pregnancy in the future, willingness for treatment was similar to willingness for future family planning (87.3% vs 83.9%), but one in five answered differently, including 10 women who intended to use contraception in future and were not willing to use it if required for treatment; their reasons could be explored only partly, because only three of the 14 non-pregnant women unwilling for treatment took part in the individual interviews. Willingness was not universal, however: 15 of the 118 women at risk (12.7%) would not accept the requirement for treatment, and almost one in three women overall (30.5%) would not join a trial requiring it, so a substantial minority could be excluded. These figures are stated willingness, not uptake. In the interviews and focus groups, acceptance was conditional on concerns about side effects, gender-related constraints, partner control and trust in the research team, and it appeared to depend more on the context in which contraception is offered than on contraception itself. Equitable access to new treatments that require contraception may therefore require more than contraceptive availability: gender-sensitive counselling, community engagement, reliable information, strengthened health-system pathways and sustained relationships of trust should be incorporated into treatment implementation and clinical research.

## Data Availability

The individual-level dataset generated during this study is not publicly available because it contains potentially identifiable sociodemographic, clinical and opinion-related information, and secondary use or external sharing of the individual-level data was not included in the original informed consent. De-identified data may be made available upon reasonable request, subject to ethical approval, applicable data-protection requirements and approval by the study’s data-access committee. Requests may be addressed to.

## Acknowledgments

We thank the participants, the community advisory board, the health teams of the participating units and the municipal health secretariats.

## Supporting information

S1 Inclusivity in Global Research: Questionnaire

S2 Checklist. Reporting checklists. STROBE (survey), SRQR (qualitative component) and GRAMMS (mixed-methods) checklists completed for this study.

S3 Human Participants checklist

S4 Interview and focus group guides. Semi-structured guides used in the in-depth interviews and in the focus groups with women community leaders, men and health professionals.

S5 Ethical approval letter

